# “Footprinting” missing epidemiological data for cervical cancer: a case study in India

**DOI:** 10.1101/2022.06.28.22276994

**Authors:** Irene Man, Damien Georges, Maxime Bonjour, Iacopo Baussano

**Author notes:** Corresponding author: Irene Man, Early Detection, Prevention and Infections Branch, International Agency for Research on Cancer, 150 cours Albert Thomas, 69372 Lyon Cedex 08, France; (IM).

## Abstract

**Background:** Context-specific cervical cancer epidemiological data are essential to derive local impact projections of cervical cancer preventive measures. However, these are not always available, in particular in low- and middle-income countries (LMICs), where impact projections are essential to plan cervical cancer control programs.

**Methods and Findings:** We developed a framework, hereafter named Footprinting, to approximate the sexual behavior, human papillomavirus (HPV) prevalence, and/or cervical cancer incidence data needed for impact projections. The framework was applied to a case study in India, the country with the highest expected cervical cancer burden but still limited access to cervical cancer prevention. With our Footprinting framework, we 1) identified clusters of Indian states with similar cervical cancer incidence patterns, 2) classified states without incidence data to the identified clusters based on similarity in sexual behavior data, 3) approximated missing cervical cancer incidence and HPV prevalence data based on available data within each cluster. Two main patterns of cervical cancer incidence, characterized by high and low incidence, were identified for 6 and 8 Indian states, respectively. States in the low-incidence cluster were characterized by less sexual activity with non-regular partners in men and earlier sexual debut in women. Based on these patterns, all 11 Indian states with missing cervical cancer incidence data were classified to the low-incidence cluster. Finally, missing data on cervical cancer incidence and HPV prevalence were approximated based on the mean of the available data within each cluster.

**Conclusions:** With the Footprinting framework, we enabled approximation of missing cervical cancer epidemiological data and derivation of context-specific impact projection of cervical cancer prevention measures, assisting public health decisions on cervical cancer prevention in India and other LMICs.

## Introduction

Cervical cancer is an important source of disease burden worldwide.[1] In 2020, the number of new cases and deaths due to cervical cancer worldwide were estimated to be 604,000 and 342,000, respectively.[2] Vaccination against human papillomavirus (HPV), cervical cancer screening, and treatment of pre-cancer and cancer can reduce the burden of cervical cancer,[3-5] but access to these prevention measures is still limited in many places of the world, especially in low- and middle-income countries (LMICs).[6, 7] To encourage the scale-up of cervical cancer prevention worldwide, the World Health Organization has developed a global strategy to eliminate cervical cancer as a public health problem.[8] The strategy proposes an elimination target of 4 cases per 100,000 women-years (age-standardized) and three intervention targets: 90% of girls vaccinated against HPV by age 15; 70% of women receiving twice-lifetime screening with high-performance testing; and 90% of women having access to cervical pre-cancer and cancer treatment, and palliative care.

In order for the aspirational global targets to be perceived as realistic, achievable, and equitable, they must be adapted to local context.[9] The local need for and the impact of cervical cancer prevention measures depend on the burden of cervical cancer in a given population, which is determined by context-specific sexual behavior, and related to the HPV prevalence.[10] Local data on these aspects are therefore crucial for deriving projections of the health and economic impact of possible interventions. When based on adequate data, impact projections of cervical cancer prevention measure can help local health authorities set adequate public health targets and allocate resources accordingly.[11]

However, local epidemiological data for cervical cancer needed to derived impact projections are sometimes missing. High-quality type- and age-specific data on HPV prevalence and cervical cancer incidence from local populations are often unavailable. The same holds for adequate data on sexual behavior, e.g., data on sex outside marriage, which are also prone to bias, e.g., social desirability and recall bias.[12, 13] When essential epidemiological data for projections are missing, there are two main possible solutions: collection or approximation of the necessary data. Collection of new data in a local context would be the ideal option. However, this could be time-and resource-demanding and therefore not always feasible. Alternatively, missing data on a given population can be approximated using available data from populations sharing similar characteristics.

In this paper, we propose a framework, hereafter named Footprinting, for approximating missing cervical cancer epidemiological data for a selected number of geographical units within a larger geographical target area where we would like to derive impact projections of cervical cancer prevention. The framework is presented through a case study in India, the country with the world’s highest expected burden of cervical cancer [5] but only very limited access to cervical cancer prevention measures.[14] To assist local public health decision-making in India, we applied Footprinting to approximate missing Indian state-specific cervical cancer incidence and HPV prevalence data and so to enable impact projections of cervical cancer preventive measures with state-specific granularity.

## Methods and Materials

### Background to the Indian case study

India, the entire target geographical area of this case study, consists of 25 geographical units of states or groups of states (both referred to as states hereafter). Detailed data on sexual behavior are available in all Indian states through a national survey.[15] However, we only identified 14 out of 25 Indian states with cervical cancer incidence data [16, 17], and two Indian states with the high-quality type- and age-specific HPV prevalence data as well as the cervical cancer incidence data needed for impact projections.[18-20] See **Table S1** for the definition of the 25 states and an overview of data availability by state.

### Footprinting framework

We propose a framework, Footprinting, to approximate missing data on the three key aspects of cervical cancer epidemiology: sexual behavior, HPV prevalence, and cervical cancer incidence. Missing data across geographical units are assumed to occur in a hierarchical manner, i.e., there is an ordering of geographical units according to their levels of data availability (**Figure 1**). At the highest level, there are a small number of geographic units for which data on all key aspects are available. For the Indian case study, there were two such states. The remaining geographic units are further divided into the levels of intermediate and low data availability. In the Indian case study, there were 12 states with cervical cancer incidence and sexual behavior data, which were assigned to the level of intermediate data availability. The remaining 11 states had only sexual behavior data and were assigned to the low level of data availability. For reason that will become clear shortly, the three data sources with increasing data availability are labelled as “Bottleneck”, “Pattern” and “Footprint” data (**Figure 1**).

**Figure 1.**
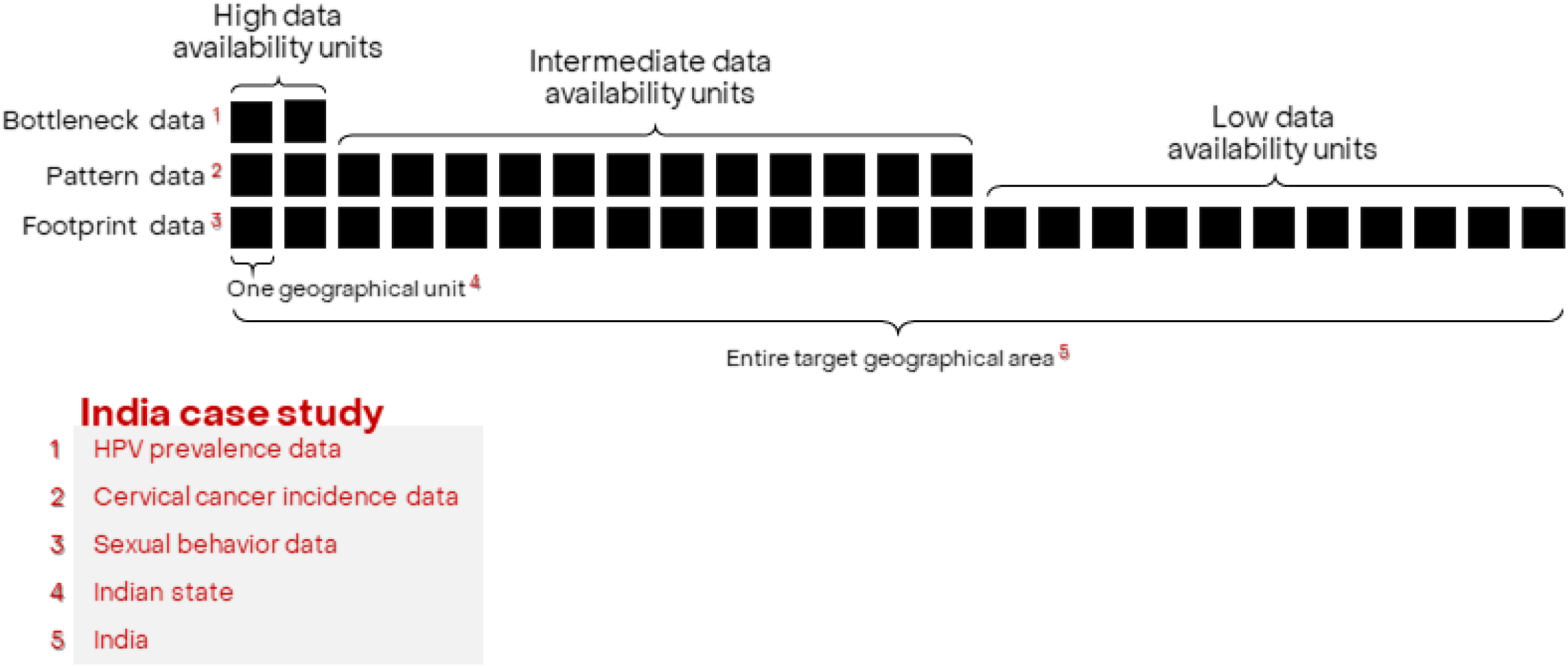
Hierarchical structure of availability of cervical cancer epidemiological data.

To address the hierarchical form of missing data, we propose a three-step approach, with the successive steps labelled as “Clustering”, “Classification”, and “Projection”. In brief, the approach identifies clusters of geographical units sharing similar patterns of cervical cancer epidemiology and uses the available data within each cluster to approximate data and extrapolate impact projections to geographical units with lower data availability. The details of the respective steps are as follows.

#### 1. Clustering step

In the Clustering step, clusters of geographical units sharing similar patterns of cancer epidemiology are identified based on the Pattern data, which has large enough coverage over the target geographical area. In the Indian case study, cervical cancer incidence data were available in 14 out of 25 Indian states and hence suitable for this purpose. As a result of the Clustering step, each Indian state of intermediate level of data availability was matched with a state with high level of data availability that shared a similar pattern of cancer incidence. In order for all states of intermediate data availability to be matched, the number of clusters must be chosen so that each cluster contained at least one state with high data availability.

#### 2. Classification step

In the second step, geographical units of the lowest level of data availability, which were not clustered in the previous step, are classified into the identified clusters. Classification is based on the similarity between geographical units according to the Footprint data, which should be available for the entire target geographical area, i.e., sexual behavior data in the Indian case study. As in the Clustering step, the result of the Classification step is that each Indian state with the lowest level of data availability was matched to states with higher levels of data availability within the same cluster.

#### 3. Projection step

In the last step, missing data are approximated based on the available data from other geographical units within the same cluster, e.g., mean or median of the available data. If the Classification step also provided the probability of belonging to each cluster, approximation could even be based on weighted values of different clusters. With the approximated data, it is then possible to construct projection models, i.e., HPV transmission and cervical cancer progression models, and derive context-specific impact projections for each geographical unit separately. Alternatively, a less computationally demanding approach would be to construct projection models for the geographical units of the highest level of data availability only and to, subsequently, scale the derived projections to the other geographical units within the same clusters.

### Data sources

In this section, we describe the data sources used in the India case study. The primary source of cervical cancer incidence data, which was used as Pattern data in the Clustering step, was cancer registry data from volume XI of Cancer Incidence in Five Continents (CI5).[16] It comprised incidence data from 16 cancer registries in 10 of the 25 Indian states (some states had more than one registry). In addition, cervical cancer incidence data were extracted from the 2012-2016 report by the Indian National Centre for Disease Informatics and Research (NCDIR) to provide data from 17 additional cancer registries not included in CI5.[17] Combining the two sources provided incidence data for 33 registries in 14 Indian states. Cervical cancer incidence was reported in number of cases per 100,000 women-years, stratified by 5-year age groups from age 15 to 79 years. See **Figure S1** for the extracted incidence data by state.

Sexual behavior data, which were used as Footprint data in the Classification step, were from the National Behavior Surveillance Survey by the National AIDS Control Organization of India in 2006, which was the most recent edition at the moment of writing.[15] Data for all 25 Indian states were available in the survey. Sexual behavior data by Indian state in the form of aggregate statistics of survey respondents were available for the following 4 groups of 12 variables:

- *Median age of first sex* – stratified by residence *(urban/rural*) and sex (*male/female*), resulting in 4 variables.
- *Proportion of respondents reporting sex with non-regular partners in the last 12 months* − stratified by residence (*urban/rural*) and sex (*male/female*), resulting in 4 variables.
- *Proportion of male respondents reporting sex with commercial partners in the last 12 months* – stratified by residence (*urban/rural*), resulting in 2 variables.
- *Proportion of male respondents by number of commercial partners in the last 12 months* − restricted to respondents with at least one commercial partner and divided into three categories (*1/2-3/>3*). As the 3 proportions always sum up to one and are therefore correlated, we omitted one category, resulting in 2 variables.

See **Figure S2** for the extracted sexual behavior data by state.

### Statistical methods

The statistical method employed in the Clustering step to cluster registry-specific cervical cancer incidence data was a Poisson-regression-based CEM clustering algorithm,[21, 22] described in detail in **Appendix S1**. Briefly, clusters of age-specific cervical cancer incidence were obtained by likelihood-based optimization under Poisson regression model. The Poisson regression model for each cluster was characterized by three parameters: an intercept, one parameter for age, and one for the square of age. This parametric form was chosen to match the general pattern of incidence through age, namely, increasing from zero incidence from the youngest age group, then decreasing after reaching a maximum (**Figure S1**). Application of the clustering method required prefixing the number of clusters *k*. The goodness-of-fit of each *k*-clustering was evaluated based on the Bayesian information criterion (BIC). To transform the obtained clustering of registry-specific data to clustering of Indian states for states with multiple registries, we assigned each state to the cluster that included the highest number of its registries, i.e., according to a majority rule.

In the Classification step, we assigned the remaining states without cervical incidence data to the identified clusters based on Random Forest (RF) using the sexual behavior data as Footprint data.[23] The RF classifier was constructed using sexual behavior data from states with identified clusters. The predictive value of each variable was evaluated with the mean decrease in accuracy, which expressed how much the accuracy of the model decreased if the variable were excluded. The performance of the classification step was validated by both out-of-bag error estimate and by applying the constructed classifier to the sexual behavior data from states with identified clusters. Subsequently, the constructed classifier was applied to the sexual behavior data from states without identified clusters, providing the probability to belong to each cluster. Each state was classified to the cluster receiving the highest probability. Classification was performed using the R package *party*. See **Appendix S2** for details of the RF-based classification method.

Finally, in the projection step, missing data on cervical cancer incidence and HPV prevalence were approximated based on the mean within each cluster. Derivation of impact projections was reported elsewhere.[24, 25]

## Results

### Clustering of cervical cancer incidence patterns

Clusterings of registry-specific cervical cancer incidence were obtained for up to four prefixed clusters (**Figure 2**). Model fit improved substantially when increasing the number of prefixed clusters from two to three, with the BIC reducing from 6933 to 5700 (**Table 1**). Further increase in the number of prefixed clusters to four only led to marginal improvement in model fit, with a small reduction in BIC from 5700 to 5532, and a poorly defined cluster of only one registry. With the number of prefixed clusters set at five, the clustering method no longer converged. We concluded that two or three clusters were fitting to describe the patterns of cervical cancer incidence in the available data.

**Figure 2.**
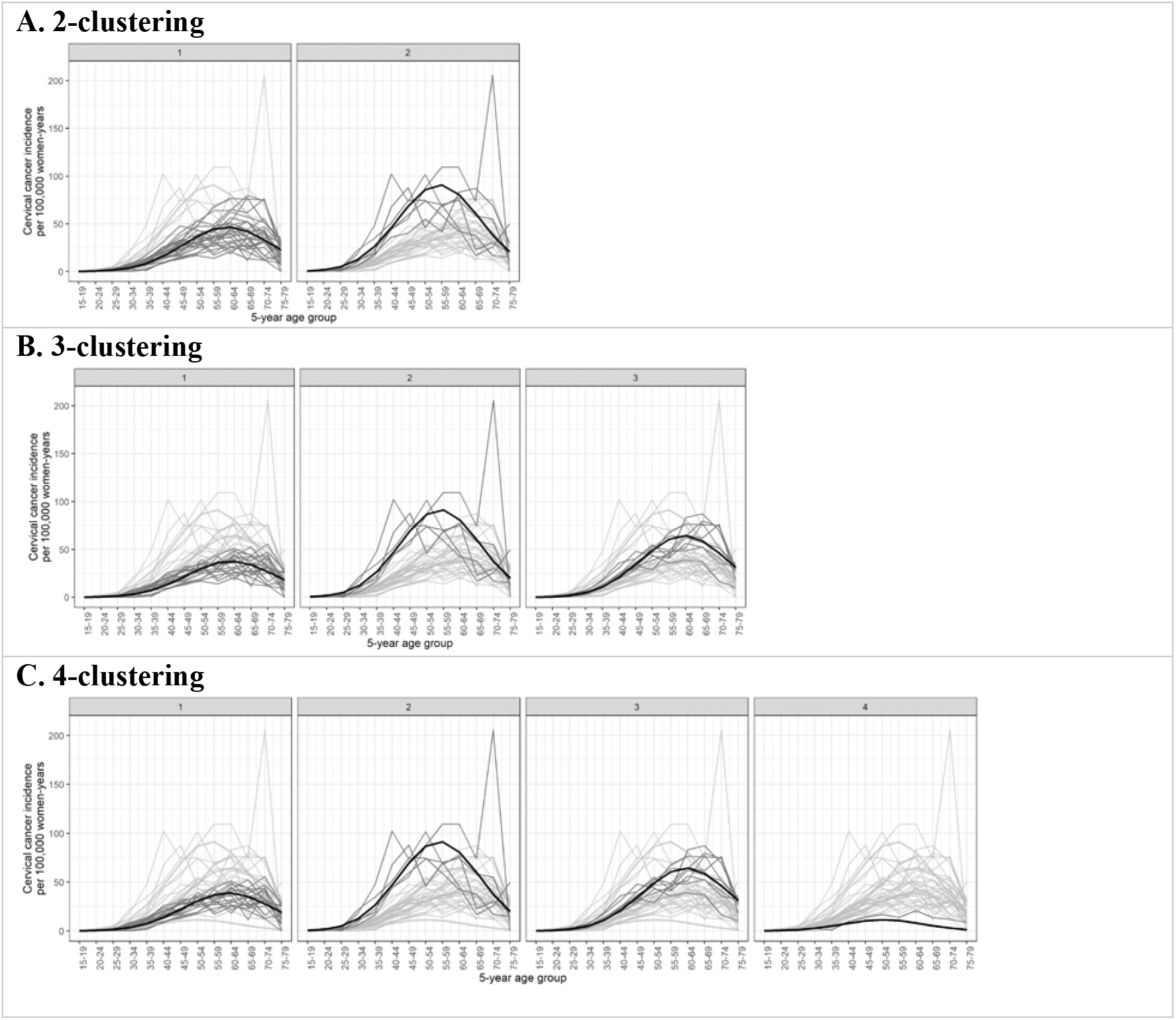
Identified clusters of registry-specific cervical cancer incidence. Clusterings under (A) 2, (B) 3, and (C) 4 pre-fixed clusters. Each panel within a row corresponds to a cluster within a *k*-clustering, with the cluster label given on top of the panel. The cervical cancer incidence data were extracted from volume XI of Cancer Incidence in Five Continents (CI5) [16] and the 2012-2016 report by the Indian National Centre for Disease Informatics and Research (NCDIR) [17]. Black: cluster mean of cervical cancer incidence; Dark gray: registry incidence assigned to the cluster; Light gray: registry incidence assigned to other clusters.

**Table 1.** Estimated parameters of clusters of cervical cancer incidence patterns.

| Number of prefixed clusters | BIC * | Cluster label *i* | Number (%) of registries in cluster | Maximum incidence § | Maximum incidence pattern | Maximum incidence age group † | Maximum incidence age group pattern |
| --- | --- | --- | --- | --- | --- | --- | --- |
| 2 | 6933 | 1 | 27 (82%) | 47 cases | Low | 60-64 year | Late |
|  |  | 2 | 6 (18%) | 91 cases | High | 55-59 year | Early |
| 3 | 5700 | 1 | 19 (58%) | 38 cases | Low | 60-64 year | Late |
|  |  | 2 | 5 (15%) | 92 cases | High | 55-59 year | Early |
|  |  | 3 | 9 (27%) | 64 cases | Intermediate | 60-64 year | Late |
| 4 | 5532 | 1 | 18 (55%) | 39 cases | Low | 60-64 year | Late |
|  |  | 2 | 5 (15%) | 92 cases | High | 55-59 year | Early |
|  |  | 3 | 9 (27%) | 64 cases | Intermediate | 60-64 year | Late |
|  |  | 4 | 1 (3%) | 20 cases | Very low | 60-64 year | Early |

\* Bayesian information criterion for evaluating the goodness-of-fit of obtained clustering
§ Maximum incidence given in cases per 100,000 women-years
† Five-year age group in which the maximum incidence is located

The identified clusters of cervical cancer incidence differed in terms of magnitude of incidence and location of maximum incidence (**Figure 2**). Cluster 1 of the 2-clustering had a low maximum incidence of 47 cases per 100,000 women-years at age group 60-64 years, compared to cluster 2 with its higher maximum incidence of 91 cases per 100,000 women-years at the earlier age group of 55-59 years (**Figure 2, Table 1**). When allowing three clusters, an additional cluster characterized by intermediate maximum incidence of 64 cases per 100,000 women-years at age group 60-64 years was identified (**Figure 2, Table 1**). This additional cluster mainly consisted of registries that had previously been assigned to the low-incidence cluster, i.e., cluster 1 of the 2-clustering, while having a relatively high incidence (**Figure 2, Table 2**). See supplementary **Table S2** for additional details of the obtained clusterings.

**Table 2.** Clustering of cervical cancer incidence of Indian states based on clustering of registries.

| State/group of states * | 2-clustering |  | 3-clustering |  |  | 4-clustering |  |  |  |
| --- | --- | --- | --- | --- | --- | --- | --- | --- | --- |
|  | 1<br>(low, late) | 2<br>(high, early) | 1<br>(low, late) | 2<br>(high, early) | 3<br>(interm., late) | 1<br>(low, late) | 2<br>(high, early) | 3<br>(interm., late) | 4<br>(very low, early) |
| Andhra Pradesh | • |  | • |  |  | • |  |  |  |
| Assam | ••• |  | ••• |  |  | •• |  |  | • |
| Delhi | • |  |  |  | • |  |  | • |  |
| Gujarat + Dadra & Nagar Haveli | • |  | • |  |  | • |  |  |  |
| Karnataka | • |  |  |  | • |  |  | • |  |
| Kerala + Lakshadweep | •• |  | •• |  |  | •• |  |  |  |
| Madhya Pradesh | • |  |  |  | • |  |  | • |  |
| Maharashtra | •••••• | • | •••• |  | ••• | •••• |  | ••• |  |
| Manipur | •• |  | •• |  |  | •• |  |  |  |
| Other North Eastern States § | ••••• | •••• | •••• | •••• | • | •••• | •••• | • |  |
| Punjab + Chandigarh | • |  |  |  | • |  |  | • |  |
| Sikkim | • |  | • |  |  | • |  |  |  |
| Tamil Nadu + Puducherry | • | • |  | • | • |  | • | • |  |
| West Bengal + Andaman & Nicobar Islands | • |  | • |  |  | • |  |  |  |

Each circle represents the count of one registry being assigned to the corresponding cluster. Gray shading represents the cluster including the highest number of registries, either exclusively or in a draw with another cluster.
\* States/or groups of states were defined as reported in the 2006 National Behavior Surveillance Survey of the National AIDS Control Organization of India.[15]
§ Other North Eastern States included Arunachal Pradesh, Nagaland, Meghalaya, Mizoram, and Tripura.
Abbreviation: Interm, intermediate.

The obtained clusterings of registries were then used to derive clusterings of Indian states based on the majority rule (**Table 2**). When using the 2-clustering of registries, none of the states were exclusively attributed to cluster 2, hence 2-clustering could not be used for the classification step. When using 3-clustering, still none of the states were exclusively attributed to cluster 2, however, 8 and 4 states were assigned to clusters 1 and 3, i.e., the clusters with low and intermediate incidence, respectively. Hence, we combined cluster 2 and 3, i.e., the clusters with high and intermediate incidence as the new “high-incidence” cluster, while keeping cluster 1 as the “low-incidence” cluster. Note that, with these newly defined clusters, each cluster still contained at least one state with the highest level of data availability, which was necessary for the projection step. Note, however, that with the new definition of clusters, we could no longer distinguish patterns of early or late peak of incidence.

### Classification to cervical cancer patterns based on sexual behavior data

A RF classifier was constructed using the sexual behavior data corresponding to the states with identified clusters. The variables with the highest, second, and third highest predictive values were *“proportion of urban male respondents reporting sex with non-regular partners in the last 12 months”, “median age of first sex in rural males”*, and *“median age of first sex in urban females”*, respectively (**Table S3**). In particular, there was a good separation between the high- and low-incidence clusters in terms of *“proportion of urban male respondents reporting sex with non-regular partners in the last 12 months”*, with high proportions associated with the high-incidence cluster (**Figure 3**). High values of *“median age of first sex in females”* and low values of *“median age of first age in males”* were also associated for the high-incidence cluster, although the separation was less clear.

**Figure 3.**
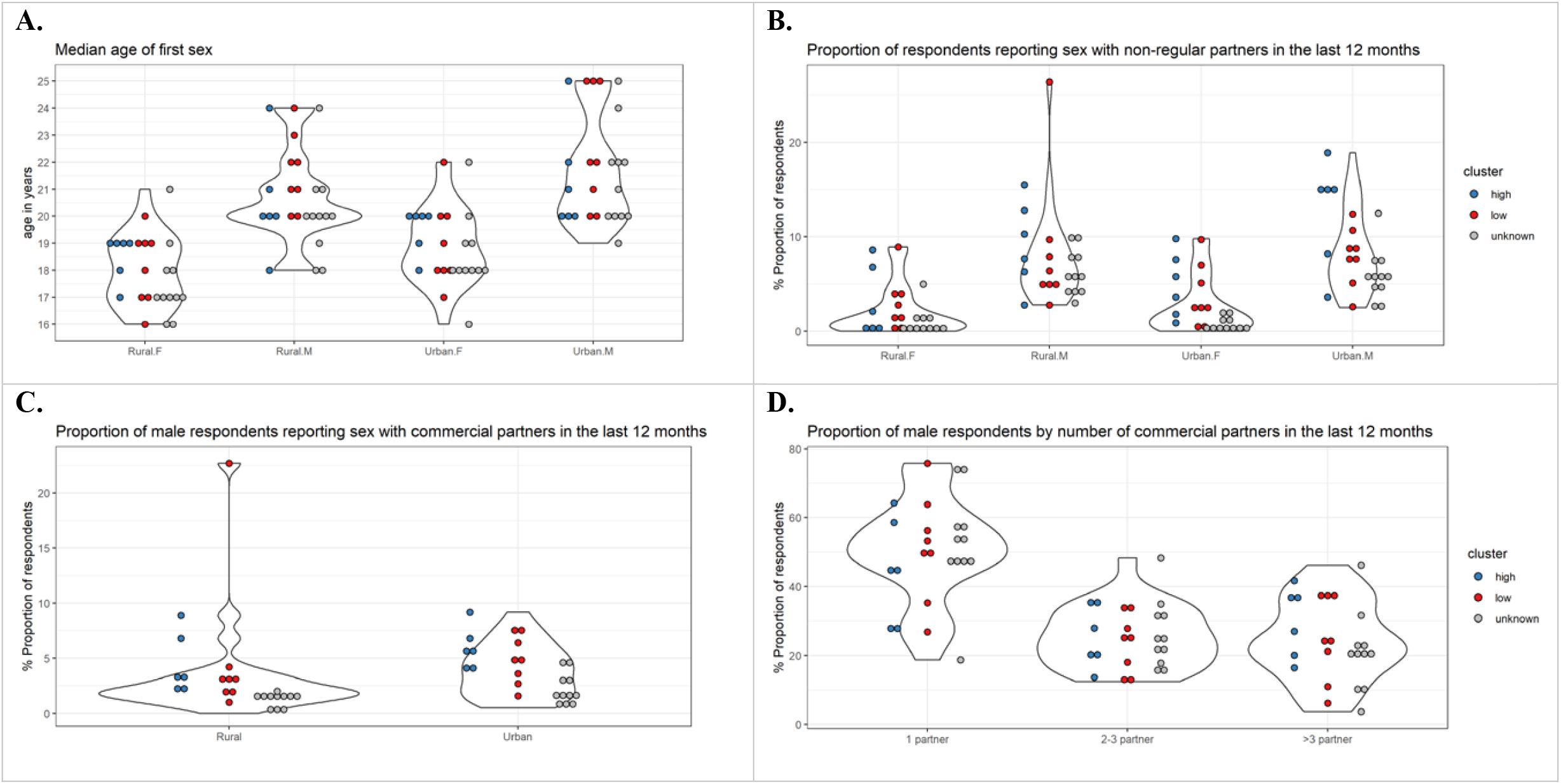
Sexual behavior data from NACO by Indian state. Indian state-specific data on (A) median age of first sex, (B) proportion of respondents reporting sex with non-regular partners in the last 12 months, (C) proportion of male respondents reporting sex with commercial partners in the last 12 months, and (D) proportion of male respondents by number of commercial partners in the last 12 months. Each violin plot and the associated cloud of circles correspond to a sexual behavior variable. Each circle corresponds to the data of a state (or group of states). The data were extracted from the 2006 National Behavior Surveillance Survey of the National AIDS Control Organization of India.[15] Blue and red: Indian states identified in the high and low cervical cancer incidence clusters. Gray: states without cervical cancer incidence data and therefore unknown cluster.

The estimated out-of-bag error of the constructed classifier was 29%. When applying the constructed classifier to the Indian states with identified clusters, only Karnataka and Other North Eastern States (2 of 14 states; 14%) were wrongly classified to the low-incidence cluster (**Table 3**). Visualization shows that the sexual behavior data of these two states more resemble the other states belonging to the low-incidence cluster, despite being clustered into the high-incidence cluster (**Figure S2, Figure 3**).

**Table 3.** Identified and classified cluster of cervical cancer incidence pattern by Indian state.

| Cervical cancer incidence data ° | State/group of states * | Identified cluster † | Classified cluster ‡ | Probability of belonging to the low-incidence cluster |
| --- | --- | --- | --- | --- |
| Available | Andhra Pradesh | low | low | 0.60 |
|  | Assam | low | low | 0.69 |
|  | Delhi | high | high | 0.42 |
|  | Gujarat + Dadra & Nagar Haveli | low | low | 0.69 |
|  | Karnataka | high | low | 0.63 |
|  | Kerala + Lakshadweep | low | low | 0.60 |
|  | Madhya Pradesh | high | high | 0.44 |
|  | Maharashtra | low | low | 0.57 |
|  | Manipur | low | low | 0.65 |
|  | Other North Eastern States§ | high | low | 0.53 |
|  | Punjab + Chandigarh | high | high | 0.41 |
|  | Sikkim | low | low | 0.63 |
|  | Tamil Nadu + Puducherry | high | high | 0.38 |
|  | West Bengal + Andaman & Nicobar Islands | low | low | 0.71 |
| Unavailable | Bihar | - | low | 0.67 |
|  | Chhattisgarh | - | low | 0.66 |
|  | Goa + Daman & Diu | - | low | 0.54 |
|  | Haryana | - | low | 0.66 |
|  | Himachal Pradesh | - | low | 0.58 |
|  | Jammu & Kashmir | - | low | 0.63 |
|  | Jharkhand | - | low | 0.71 |
|  | Orissa | - | low | 0.68 |
|  | Rajasthan | - | low | 0.66 |
|  | Uttar Pradesh | - | low | 0.64 |
|  | Uttarakhand | - | low | 0.69 |

° Availability of cervical cancer incidence data was based on the incidence data from volume XI of Cancer Incidence in Five Continents (CI5) and the 2012-2016 report of the National Centre for Disease Informatics and Research (NCDIR).[16, 17]
\* States/groups of states were defined as reported in the 2006 National Behavior Surveillance Survey of the National AIDS Control Organization of India.[15]
§ Other North Eastern States included Arunachal Pradesh, Nagaland, Meghalaya, Mizoram, and Tripura.
† Identified clusters derived in the Clustering step.
‡ Classified clusters derived in the Classification step. A given state was classified to the low-incidence cluster if the probability of belonging to the low-incidence cluster (given in the next column) was above 0.50. For the Indian states with available cervical cancer incidence data and hence already in an identified cluster, classification was done for the purpose of validation.

Subsequently, the constructed classifier was applied to classify the remaining states without cervical cancer incidence data and thus with unknown cluster. All 11 remaining Indian states received a higher probability to belong to the low-incidence cluster (**Table 3**). Indeed, **Figure 3** shows that the sexual behavior data of the states with unknown cluster (indicated in gray) were generally closer to the sexual behavior data of the states of the low-incidence cluster (indicated in red) than those of the high-incidence cluster (indicated in blue). Hence, we identified in total 19 and 6 states for the low- and high-incidence clusters, respectively.

Finally, missing cervical cancer incidence data and HPV prevalence were approximated based on the mean within each cluster (**Figure S3**). Approximation of HPV prevalence was based on the only one prevalence survey we could identify per cluster.(20,21) We verified that the HPV prevalence reported by the survey corresponding to the high-incidence cluster was higher than the prevalence reported by the one corresponding to the low-incidence cluster: HPV prevalence of 16.9% vs. 9.8% (in women in the age range of approximately 20-60 years). This 1.7-fold difference in HPV prevalence was in the same order of magnitude as the 1.9-fold difference we found for the age-standardized cervical cancer incidence between the two clusters (17.9 vs. 9.01 cases per 100,000 women-years). The final step of deriving impact projections of cervical cancer preventive measures for the whole of India with state-specific granularity was reported elsewhere.[24, 25]

## Discussion

In this paper, we developed the Footprinting framework to approximate missing cervical cancer epidemiological data in some geographical units when deriving impact projections of cervical cancer prevention measures for a larger geographical area. In brief, the framework identified clusters of geographical units sharing similar patterns of cervical cancer epidemiology and uses the available data within each cluster to approximate data and extrapolate impact projections to geographical units with lower data availability. The framework was demonstrated by a case study of approximating missing cervical cancer incidence and HPV prevalence data for a selection of Indian states. With the application, we have derived, for the first time, impact projections of cervical cancer prevention measures for the whole of India with state-specific granularity.[24, 25]

Moreover, this work has also generated a deeper understanding of cervical cancer epidemiology throughout the country. We found that India can be divided into two main groups of 19 and 6 Indian states or groups of states that are characterized by low or high cervical cancer incidence, respectively. As expected, and in line with previously studies, individuals, in particular men, in high-incidence states have more sexual activity with non-regular partners, including commercial partners, than in low-incidence states.[26, 27] While early sexual debut in women has also previously been suggested to be associated with high cervical cancer incidence and HPV positivity,[26, 27] it was associated with lower cervical cancer incidence in the dataset we considered. We hypothesize that, for the Indian context, early sexual debut is common in states with a larger rural population, among whom less sexual activity occurs with non-regular partners, which is the main determining factor for a lower risk of cervical cancer. With the urbanization of rural areas, which often entails evolving socio-cultural norms, it is possible that more Indian states may shift to a high cancer incidence pattern, with an accompanying early peak in incidence.[28]

It should be noted that our Footprinting framework is similar to other extrapolation approaches previously used in model-based projection studies targeting large geographical areas with missing data.[29, 30] While the rationale behind different extrapolation approaches are similar, which is to approximate missing data from other geographical units with similar epidemiological indicators, there are also differences. A strength of our framework is that it relies on the observed patterns of epidemiology in the data to select key geographical units from which impact projections are extrapolated to other units instead of working with a predefined selection of key units. This allows the selection of key units that maximizes the representation of different epidemiological patterns in the data. Moreover, it also helps to pinpoint geographical units that could be interesting for future data collection efforts. Secondly, we made use of a newly developed clustering method [21, 22] that is able to assess the similarities between age-specific patterns of cervical cancer incidence, which have not been considered by previous studies.

Our application of Footprinting to the Indian case study also bears some resemblance with the extrapolation of cervical cancer incidence by GLOBOCAN in the process to obtain nationwide estimates of cervical cancer incidence in India.[31] Essentially, in GLOBOCAN, missing incidence was extrapolated based on the identity being urban or rural as a footprint, while we used sexual behavior for this purpose and considered each state separately, which is necessary for state-specific impact projections. As a result, we neglected the variation between rural and urban areas within Indian states, which is a limitation of our analysis. We expect that Footprinting with further stratification of states by rural/urban identity could improve the approximation. Furthermore, our nationwide estimate of cervical cancer incidence derived from aggregating the state-specific estimates (reported in a separate manuscript [24]) was lower than the estimate reported by GLOBOCAN. This could be explained by the use of different methods of extrapolation and the fact that we included data from 17 additional cancer registries with relatively low incidence not included in GLOBOCAN estimates.

Various possible adaptations of the proposed Footprinting framework are worth mentioning. Firstly, in the less than ideal situation where none of the relevant cervical cancer epidemiology data are available in some geographical units, data on indicators of human development and geographical location, could also be used as Footprint data. Secondly, while we focused on epidemiological data for cervical cancer in this work, Footprinting could be used to approximate missing economic data (e.g., treatment or vaccine delivery costs) that are needed to assess the health economic impact of cervical cancer prevention measures, given that relevant footprint data can be defined and collected.

Through this work, we have provided a comprehensive framework to deal with the important and ubiquitous challenge of missing data on cervical cancer epidemiology. By using the proposed framework, it is possible to derive robust yet context-specific impact projections for cervical cancer preventive measures for a wide range of geographical settings. Such projections can assist local health authorities in planning and implementing cervical cancer prevention that is adapted to the local needs and resources and so intensify the efforts to reduce the high burden of cervical cancer still existing in many countries in low-resource settings.

## Data Availability

All data used in the present study were openly available at: https://www.aidsdatahub.org/sites/default/files/resource/national-bss-general-population-india-2006.pdf for the sexual behavior data published by the National AIDS Control Organisation Ministry of Health and Family Welfare Government of India, at http://ci5.iarc.fr for the cervical cancer incidence data published by the International Agency for Research on Cancer, and at https://www.ncdirindia.org/All_Reports/Report_2020/resources/NCRP_2020_2012_16.pdf for the cervical cancer incidence data published by the National Centre for Disease Informatics and Research of India.
All data produced in the present study are available upon reasonable request to the authors.

http://ci5.iarc.fr

https://www.ncdirindia.org/All_Reports/Report_2020/resources/NCRP_2020_2012_16.pdf

## Acknowledgments

This study was funded by the Bill & Melinda Gates Foundation (grant numbers: OPP48979; INV-039876). The funder had no role in study design, data collection and analysis, decision to publish, or preparation of the manuscript. For the authors identified as personnel of the International Agency for Research on Cancer or World Health Organization, the authors alone are responsible for the views expressed in this article and they do not necessarily represent the decisions, policy or views of the International Agency for Research on Cancer or World Health Organization. The designations used and the presentation of the material in this Article do not imply the expression of any opinion whatsoever on the part of WHO and the IARC about the legal status of any country, territory, city, or area, or of its authorities, or concerning the delimitation of its frontiers or boundaries.

## Contributors

IB was responsible for funding acquisition and supervision of this study. IM, DG, and IB co-designed and co-led the data curation, formal analysis, investigation, and visualization of the study finding. MB contributed to the development of statistical methodology and validation of study results. All authors had full access to all the data in the study and had final responsibility for the decision to submit for publication. IM and IB drafted the original draft. All authors contributed to review and editing and have approved the final manuscript.

## Data Availability Statement

All data used in the present study were openly available at: https://www.aidsdatahub.org/sites/default/files/resource/national-bss-general-population-india-2006.pdf for the sexual behavior data published by the National AIDS Control Organisation Ministry of Health and Family Welfare Government of India, at http://ci5.iarc.fr for the cervical cancer incidence data published by the International Agency for Research on Cancer, and at https://www.ncdirindia.org/All_Reports/Report_2020/resources/NCRP_2020_2012_16.pdf for the cervical cancer incidence data published by the National Centre for Disease Informatics and Research of India. All data produced in the present study are available upon reasonable request to the authors.

