## Supplementary material for "“Footprinting” missing epidemiological data for cervical cancer: a case study in India"

### List of tables

Table S1. Overview of availability of cervical cancer epidemiological data by state
Table S2. Estimated model parameters under Poisson regression models.
Table S3. Predictive values of the sexual behavior variables for cervical cancer incidence cluster

#### List of figures

Figure S1. Registry-specific cervical cancer incidence data from CI5 and NCDIR
Figure S2. Indian state-specific sexual behavior data from NACO
Figure S3. Mean age-specific cervical cancer incidence by cluster

### Appendix S1. Poisson-regression-based CEM clustering algorithm

The clustering algorithm employed to cluster registry-specific cervical cancer incidence data, which are count data, is based on a previously published model-based clustering algorithm.(1,2) The method relies on an iterative process to obtain an optimal clustering based on likelihood under a Poisson regression model. The algorithm operates with a prefixed number of clusters *k*. The iterative process is initialized by an initial C-step and an initial M-step:

- Initial C-step: randomly assign the age-specific cervical cancer incidence $Y_r$ of each registry *r* to one of the *k* clusters.
- Initial M-step: estimate the initial parameters $\theta^0 = (\beta^0_{i,intercept}, \beta^0_{i,age}, \beta^0_{i,age2})$ under the Poisson regression model as defined in Box 1 based on the initial assignment and compute the proportion of registries $\pi^0_c$ belonging to each cluster $c$.

Upon initialization, the *m*-th iteration consists of an E-step, C-step, and M-step:

- E-step: compute the probability for the age-specific cervical cancer incidence $Y_r$ of each registry $r$ to belong to each cluster $c$ given the previously obtained parameter estimates $\theta^{m-1}$:
$$p^m_{r,c} = \frac{\pi^{m-1}_c f(Y_r, \theta^{m-1}_c)}{\sum^k_{c\prime} \pi^{m-1}_{c\prime} f(Y_r, \theta^{m-1}_{c\prime})}.$$
- C-step: assign each the age-specific cervical cancer incidence of each registry to the cluster with the maximum probability.
- M-step: estimate the parameters $\theta^m = (\beta^m_{i,intercept}, \beta^m_{i,age}, \beta^m_{i,age2})$ under the Poisson regression model as defined in Box (1) based on the assignment obtained in the C-step and compute the proportion of registries $\pi^m_c$ belonging to each cluster $c$.

The iterative process terminates when the model fit no longer improves, which is defined as when the difference between two consecutive log-likelihoods is smaller than the given threshold value (0.001).

As different initial assignments could result in different final assignments, the above iterative process was repeated 100 times with different initial assignments. The 100 different final assignments were compared based on the Bayesian information criterion (BIC), and only the one with the highest BIC was selected.

**Box 1. Poisson regression model defined for the clustering algorithm above.**

A Poisson regression model with cluster-specific effects for intercept, age group id $j$ ($= 1, \ldots, 13$ for 5-year age groups 15-19, …, 75-79) and the square of age group id $j^2$ is fitted to the cervical cancer incidence data ($Y$and $t$ for the number of cases and women-years in the data) estimated based on the classified cluster obtained:

$$\log(E(Y|j)/t) = \begin{cases} \beta_{1,intercept} + \beta_{1,age} \cdot j + \beta_{1,age2} \cdot j^2, \text{for cluster } i = 1, \\ (\beta_{1,intercept} + \beta_{i,intercept}) + (\beta_{1,age} + \beta_{i,age}) \cdot j + (\beta_{1,age2} + \beta_{i,age2}) \cdot j^2, \text{for cluster } i > 1. \end{cases}$$

For notational convenience, we introduced the following auxiliary coefficients for $* = intercept, age, age2$:

$$\gamma_{i,*} = \begin{cases} \beta_{1,*}, & \text{for cluster } i = 1, \\ \beta_{1,*} + \beta_{i,*}, & \text{for cluster } i > 1. \end{cases}$$

To facilitate interpretation of the cluster patterns, we then computed the maximum incidence $y_{i,max}$ and the 5-year age group of maximum incidences $j_{i,max}$, as follows:

$$y_{i,max} = \exp\left(\gamma_{i,intercept} - \frac{{\gamma_{i,age}}^2}{4 \cdot \gamma_{i,age2}}\right) \cdot 100000, \text{ and } j_{i,max} = \left|\frac{-\gamma_{i,age}}{2 \cdot \gamma_{i,age2}}\right|, \text{ for cluster } i.$$

### Appendix S2. Random Forest classification

Random Forest classification was performed using the R package *party* version 1.3-7 with the following setting: *cforest_control(teststat = 'quad', testtype = 'Univariate', mincriterion = 0.9, ntree = 50000, mtry = 3, maxdepth = 2, minsplit = 0, minbucket = 0).*

#### Appendix S3. Supplementary tables

**Table S1. Overview of availability of cervical cancer epidemiological data by state.**

| State/Group of states * | Sexual behaviour | Cervical cancer incidence ± | HPV prevalence † | Data availability level | CI5 registry ¤ | NCDIR registry ¤ |
|---|---|---|---|---|---|---|
| Andhra Pradesh | X | X | | Intermediate | | *Hyderabad district* |
| Assam | X | X | | Intermediate | Cachar, Kamrup Urban District | Cachar district, *Dibrugarh district*, Kamrup urban |
| Bihar | X | | | Low | | |
| Chhattisgarh | X | | | Low | | |
| Delhi | X | X | | Intermediate | | *Delhi* |
| Goa + Daman & Diu | X | | | Low | | |
| Gujarat + Dadra & Nagar Haveli | X | X | | Intermediate | Ahmedabad | Ahmedabad urban |
| Haryana | X | | | Low | | |
| Himachal Pradesh | X | | | Low | | |
| Jammu & Kashmir | X | | | Low | | |
| Jharkhand | X | | | Low | | |
| Karnataka | X | X | | Intermediate | Bangalore | Bangalore |
| Kerala + Lakshadweep | X | X | | Intermediate | Kollam, Trivandrum | Kollam district, Thi'puram district |
| Madhya Pradesh | X | X | | Intermediate | Bhopal | Bhopal |
| Maharashtra | X | X | | Intermediate | Barshi & Paranda & Bhum, Mumbai, Poona, Wardha | *Aurangabad, Osamanabad & Beed*, Barshi rural, Mumbai, Pune, Wardha district, *Nagpur* |
| Manipur | X | X | | Intermediate | | *Manipur state, Imphal West district* |
| Orissa | X | | | Low | | |
| Other North Eastern States § | X | X | | Intermediate | Mizoram, Tripura | Mizoram state, *Aizawl district*, Tripura state, *West Arunachal, Papumpare district, Meghalaya, East Khasi Hills district, Nagaland, Pasighat* |
| Punjab + Chandigarh | X | X | | Intermediate | | *Patiala district* |
| Rajasthan | X | | | Low | | |
| Sikkim | X | X | | Intermediate | Sikkim State | Sikkim state |
| Tamil Nadu + Puducherry | X | X | X | High | Chennai, *Dindigul Ambilikkai* | Chennai |
| Uttar Pradesh | X | | | Low | | |
| Uttarakhand | X | | | Low | | |
| West Bengal + Andaman & Nicobar Islands | X | X | X | High | | *Kolkata* |

* States or groups of states as reported in the 2006 National Behavior Surveillance Survey of the National AIDS Control Organization of India.(3)
§ Other North Eastern States include Arunachal Pradesh, Nagaland, Meghalaya, Mizoram, and Tripura.
± States with age-specific cervical cancer incidence data from volume XI of Cancer Incidence in Five Continents (CI5) and the 2012-2016 report of the National Centre for Disease Informatics and Research (NCDIR).(4,5)
† Type- and age-specific HPV prevalence data.(6,7)
¤ The eighteen registries CI5 and NCDIR do not have in common are in *italics*.

**Table S2. Estimated model parameters under Poisson regression models.**

| **Number of prefixed clusters** | **Cluster label $\boldsymbol{i}$** | $\boldsymbol{\gamma_{i,intercept}}$ | $\boldsymbol{\gamma_{i,age}}$ | $\boldsymbol{\gamma_{i,age2}}$ |
|---|---|---|---|---|
| 2 | 1 | -14.70 | 1.43 | -0.073 |
| | 2 | -13.64 | 1.50 | -0.085 |
| 3 | 1 | -14.71 | 1.39 | -0.071 |
| | 2 | -13.66 | 1.52 | -0.086 |
| | 3 | -14.65 | 1.48 | -0.075 |
| 4 | 1 | -14.66 | 1.39 | -0.071 |
| | 2 | -13.66 | 1.52 | -0.086 |
| | 3 | -14.65 | 1.48 | -0.075 |
| | 4 | -16.16 | 1.62 | -0.086 |

The definition of $\gamma_{i,*}$ is given Appendix S1.

**Table S3. Predictive values of the sexual behavior variables for cervical cancer incidence cluster.**

| Sexual behavior variable | Predictive value | Ranking (from high to low predictive value) |
|---|---|---|
| age.first.sex.Urban.M | -0.0042 | 7 |
| age.first.sex.Urban.F | -0.0031 | 3 |
| age.first.sex.Rural.M | 0.0001 | 2 |
| age.first.sex.Rural.F | -0.0044 | 9 |
| perc.non.regular.Urban.M | 0.0220 | 1 |
| perc.non.regular.Urban.F | -0.0045 | 10 |
| perc.non.regular.Rural.M | -0.0042 | 8 |
| perc.non.regular.Rural.F | -0.0039 | 6 |
| perc.commericial.Urban | -0.0038 | 5 |
| perc.commericial.Rural | -0.0051 | 11 |
| number.commericial.1 | -0.0033 | 4 |
| number.commericial.>3 | -0.0058 | 12 |

Predictive value is defined as the mean decrease in accuracy, which expresses how much the accuracy of the model would decrease if the variable were to be excluded. Higher values correspond to higher predictive values.

#### Appendix S4. Supplementary figures

**Figure S1. Registry-specific cervical cancer incidence data from CI5 and NCDIR.**
See Table S1 for whether registries belong to CI5 or NCDIR.(4,5)

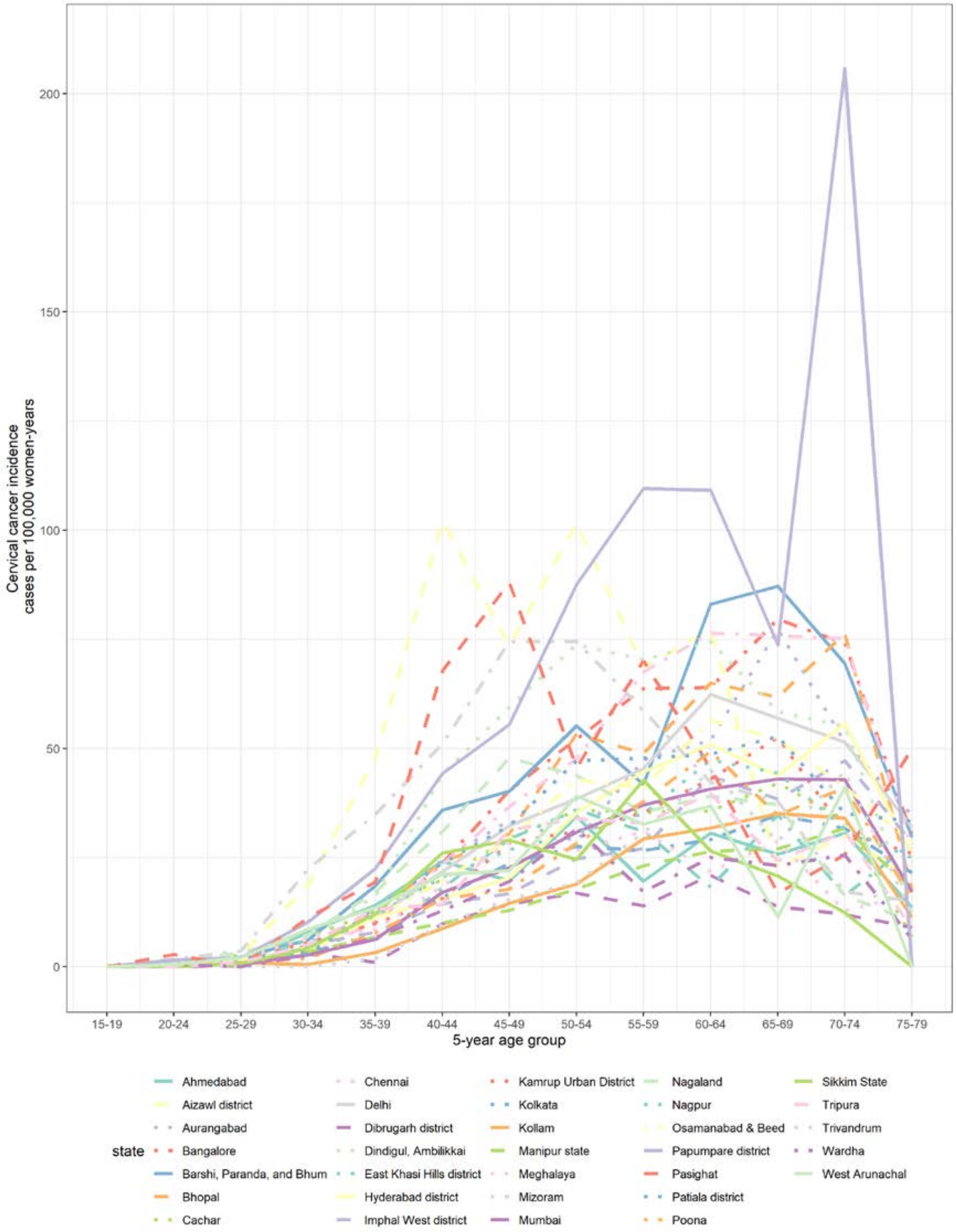

**Figure S2. Indian state-specific sexual behavior data from NACO.**
States or groups of states as reported in the 2006 National Behavior Surveillance Survey of the National AIDS Control Organization of India.(3) Other North Eastern States include Arunachal Pradesh, Nagaland, Meghalaya, Mizoram, and Tripura.

**A**

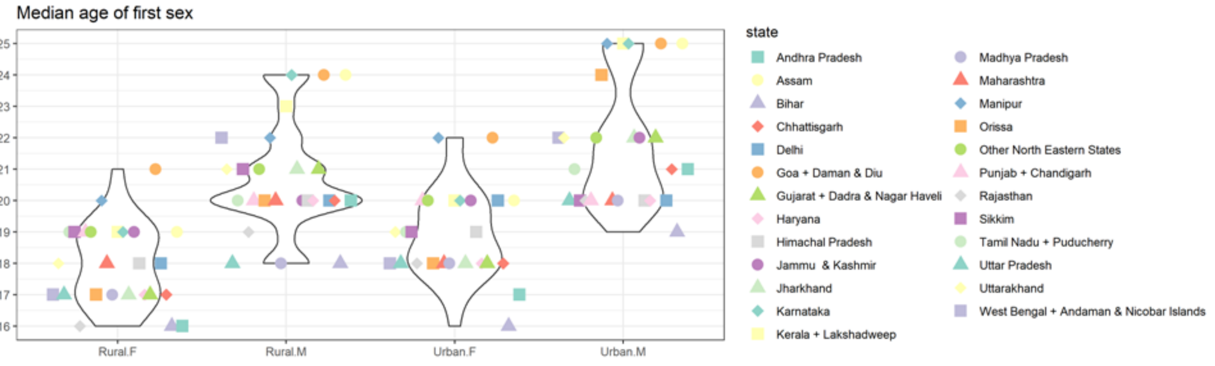

**B**

**C**

**D**

**Figure S3.** **Mean age-specific cervical cancer incidence by cluster.**

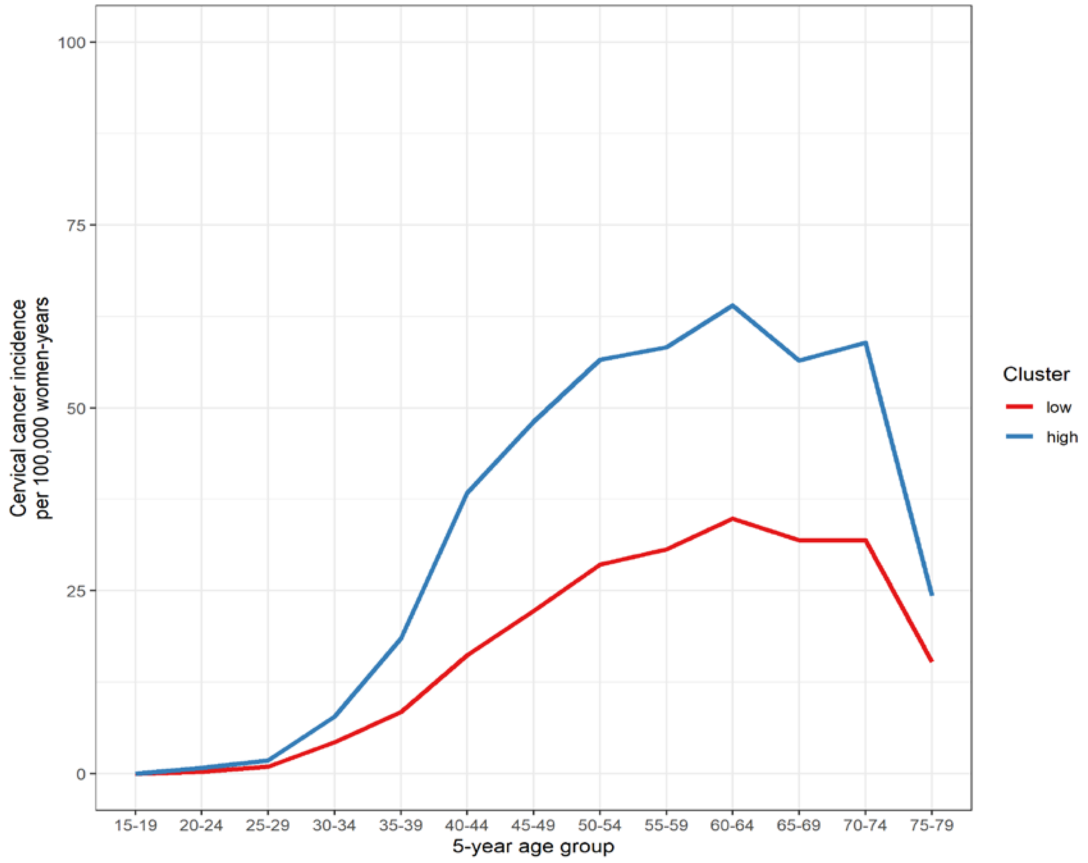
